# Echocardiographic assessment of elevated pulmonary vascular pressures after resolution of critical illness

**DOI:** 10.1101/2025.09.22.25336345

**Authors:** Andrew Wodarcyk, Sara Folk, Kali McKnight, Brian O’Connor, Scott Visovatti, Rebecca Vanderpool

## Abstract

**Background:** Pulmonary vascular pressures are freq[ABS]suently elevated in critically ill patients and are associated with worse outcomes. However, whether elevated pressures persist and their impact on outcomes after critical illness is unknown.

**Research Questions:** What factors asre associated with persistently elevated pulmonary vascular pressures? What are the outcomes associated with persistently elevated pulmonary vascular pressures?

**Study Design and Methods:** This is a single center retrospective cohort study of critically ill patients during the year 2021. Adult patients with a measured tricuspid regurgitant velocity≥2.8 m/s during critical illness and had a repeat echocardiogram done after hospital discharge were included. Kaplan Meier and logistic regression were used for mortality and multivariate analysis.

**Results:** Of 540 patients, 257 (47.6%) had an elevated TRV. Of 51 patients with a repeat echocardiogram, 33 (64.7%) had an elevated repeat TRV. These patients had higher heart rates (91±23 vs 73±14 bpm, *p<*0.01), lower hemoglobin levels (8.42±1.76 vs 10.0±2.16, *p=*0.02), decreased TAPSE (1.90 ± 0.52 mm vs 2.23 ± 0.43, p = 0.03), increased RV middle diameter(3.27±0.85 vs 2.72±0.78, *p=*0.04) and decreased left ventricular stroke volume (61.76±15.10 vs 84.36±27.09, *p=*0.01) compared to those with a normal repeat TRV. Hemoglobin (p=0.03, 95% CI: 0.30–0.90) and SVI (p=0.03, 95% CI: 0.77–0.98) were associated with elevated repeat TRV levels. Elevated TRV on repeat echocardiogram was not associated with worse survival (log-rank test, p=0.33).

**Interpretation:** Elevated pulmonary vascular pressures persisted after critical illness in a large number of patients, although the impact of persistently elevated pulmonary vascular pressures is uncertain.

## INTRODUCTION

Pulmonary vascular pressures are often elevated in critically ill patients when measured by echocardiography or right heart catheterization^1,2^. Patients in an intensive care setting with elevated pulmonary vascular pressures consistently have worse short- and long-term outcomes including increased mortality^1^. Elevated pulmonary vascular pressures can lead to increased right ventricular strain, development of intracardiac shunts and hypoxemia, and development of right ventricular failure. While the development of elevated pulmonary vascular pressures during critical illness, and the poor outcomes associated with its development are well documented, it is less known whether pulmonary vascular pressures normalize after the resolution of critical illness.

Mechanisms contributing to increased pulmonary pressure in critically ill patients are poorly understood and likely multifactorial depending on the underlying disease state.^3–7^ . In acute respiratory distress syndrome, the effects of hypoxic vasoconstriction, endothelial dysfunction, small pulmonary vessel thrombosis and obliteration, and vascular remodeling have been shown to contribute^3,4^. While in sepsis, endothelial dysfunction and small vessel thrombosis also occur but additional contributions from vasoconstrictive effects of vasopressors and bacterial endotoxins have been identified as major factors leading to increased pulmonary pressure ^5–7^. Endothelial dysfunction, small pulmonary vessel thrombosis and obliteration, and vascular remodeling are also characteristic of the pathophysiology of pulmonary arterial hypertension (PAH).^8^

Given the similarities in pathogenesis between PAH and elevated pulmonary vascular pressures in the critically ill, we hypothesize that elevated pulmonary vascular pressures would persist despite the resolution of critical illness and could be an area to explore regarding treatment with pulmonary vasodilators. In this study, we investigated whether critically ill patients with elevated pulmonary vascular pressures continue to have elevated pulmonary vascular pressures after resolution of critical illness, factors associated with persistent elevated pulmonary vascular pressures, and long-term outcomes in patients with persistently elevated pulmonary vascular pressures.

## METHODS

### Study Design

In this single-center, retrospective cohort study, patients admitted to intensive care units with an available echocardiogram at The Ohio State University Wexner Medical Center between 01/01/2021 – 12/31/2021 were identified (**Figure 1**). Echocardiographic measurements were extracted and patients with a tricuspid regurgitant velocity (TRV) measurement during their hospital stay (index echo) were included in the study. Patients less than 18 years old or with a prior diagnosis of pulmonary hypertension were excluded. Patients with a TRV≥ 2.8 at their index echocardiogram were assigned to the elevated pulmonary pressure group (TRV≥ 2.8 group). Clinical data and follow-up echocardiographic data was collected for the patients in the TRV≥ 2.8 group. The Institutional Review Board approved this study (IRB number: 2023H0288).

**Figure 1.**
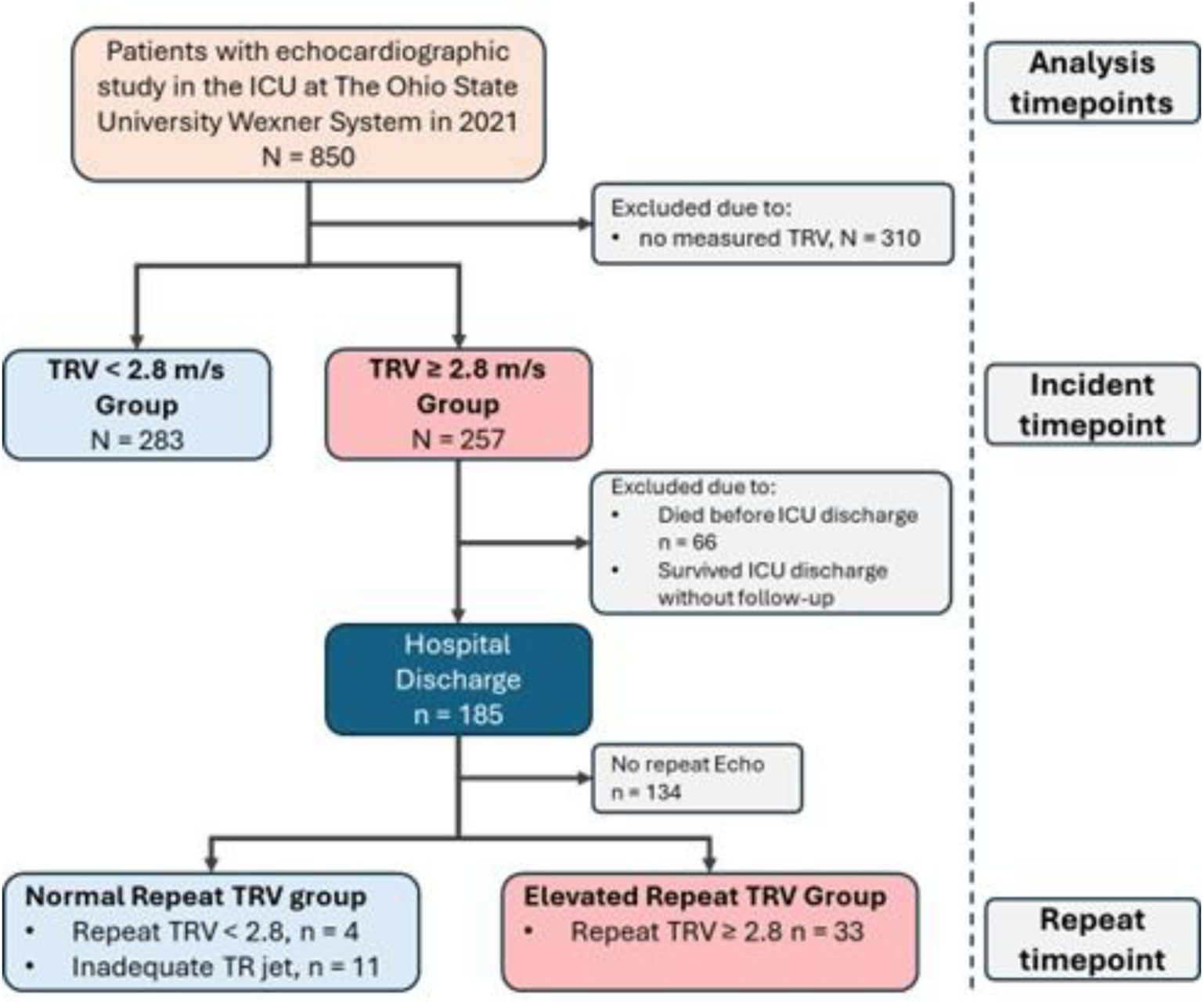
Patient Flowsheet. See table 1 legend for expansion of abbreviations.

**Table 1.**
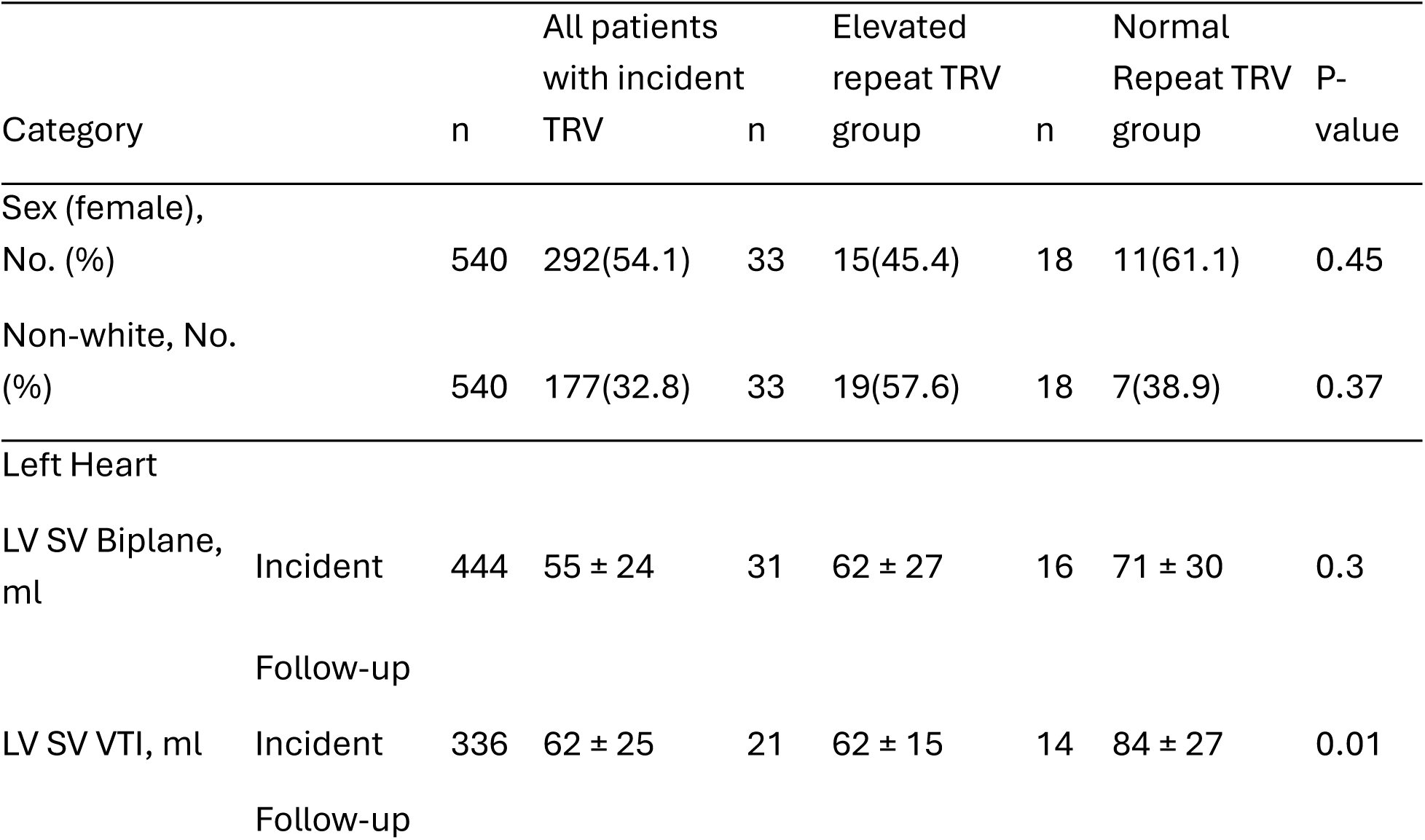
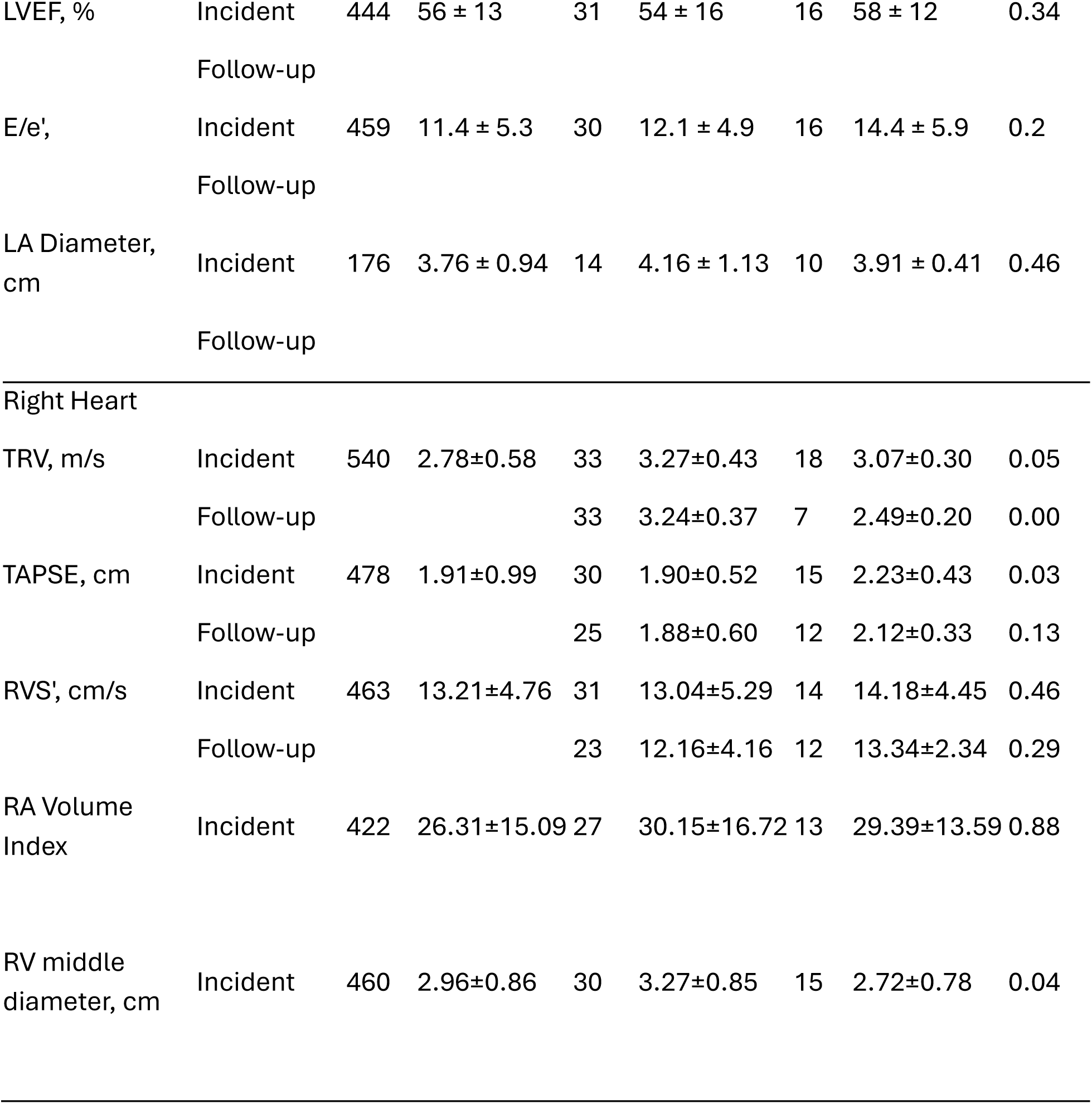
HFrEF = heart failure with reduced ejection fraction; HFpEF = heart failure with preserved ejection fraction; ARDS = acute respiratory distress syndrome; TRV = tricuspid regurgitant peak velocity; TAPSE = tricuspid annular plane systolic excursion; RVS’ = tricuspid annular velocity; LV SV Biplane = left ventricular stroke volume by biplane method; LV SV VTI = left ventricular stroke volume by velocity time integral method; LVEF = left ventricular ejection fraction; E/e’ = peak early mitral inflow velocity/early diastolic mitral annular velocity; LA = left atrium; RA = right atrium; RV = right ventricle

### Data Collection

Chart-reviews were performed on in patients with a TRV≥ 2.8 group. Data collected at the time of hospitalization included echocardiogram findings, vital signs, and results from right heart catheterization (RHC), if performed, as well as ICU duration and admitting diagnosis. Information on the use of mechanical ventilation, diuretics, inotropes, vasopressors, and pulmonary vasodilators was also recorded. Age, past medical history, admitting diagnoses, vital signs on admission, pH, sodium, potassium, creatinine, hematocrit, white blood cell count, and FiO2 were used to calculate an APACHE II score.^9^ The Glasgow Coma Scale was omitted from the APACHE II score calculation as this information was not readily accessible^9^. Baseline demographic and clinical data, including age, sex, race, comorbidities, brain natriuretic peptide (BNP), and troponin levels, were obtained.

Following hospitalization, all echocardiograms performed between ICU admission and time of collection were collected and analyzed. Patients were subsequently followed to assess for the development of pulmonary hypertension, mortality, hospital readmission, or receipt of lung transplantation.

### Statistical Analysis

Continuous variables were presented as mean ± standard deviation. Time based variables were presented as median ± interquartile range. Categorical variables were presented as numbers (n) and percentage (%). Two-tailed t-test were used to compare between groups andpaired t-tests were used to compare repeat measurements. Kaplan Meier survival analysis was used to investigate the association of elevated TRV measurements with all-cause mortality in the whole cohort. Survival time was the time between date of the incident echocardiogram and death. In the subset of patients with elevated TRV and repeat echocardiography, Kaplan Meier survival analysis was used to investigate the association of a sustained elevation in TRV with all-cause mortality. Survival time was time between date of follow-up echocardiogram and death. Survival analysis was performed using Kaplan-Meier survival with Log-Rank testing Statistical significance will be considered when p< 0.05. Univariate logistic regression analysis was used to identify baseline clinical and echocardiographic variables that are associated with elevated TRV on repeat echocardiography. Baseline variables with a p<0.5 in the univariate analysis were combined in a multivariate analysis. Data analysis was performed using R (4.1.1, R-Foundation) and Excel (Version 16.98, Microsoft Excel).

## RESULTS

### Patient characteristics

A total of 850 patients were identified to have had an echocardiographic study while in the ICU in The Ohio State University Wexner system in 2021 and were reviewed. In this initial cohort, 540 patients had a measured TRV and 257 (47.6%) had an elevated TRV (TRV ≥2.8 m/s). In the group with elevated TRV, 51 had follow-up echocardiograms post-ICU discharge. At follow-up, thirty-three patients (64.7%) had a repeat TRV ≥2.8 m/s (elevated repeat TRV group) and 18 patients had a repeat TRV <2.8 m/s or inadequate TR jet (normal repeat TRV group) (**Figure 1**).

### Baseline characteristics in patients with repeat echocardiography assessment

Baseline characteristics including sex, race, age were not significantly different between groups (Table 1). Among clinical characteristics, the elevated TRV group had an increased number of patients with pneumonia as the admitting diagnosis compared to the normal TRV group (11 patients (33.3%) vs 1 (5.6%), respectively), although this did not reach statistical significance. p=0.05). Patients in the elevated TRV group also had an elevated heart rate on ICU admission (91±23 vs 73±14 bpm, *p<*0.01) compared to the normal TRV group. All other clinical characteristics including vital signs, past medical history, admitting diagnoses, ICU duration, use and duration of vasopressors and mechanical ventilation, were not significantly different between groups. Among laboratory variables, patients in the elevated repeat TRV group had significantly increased BNP levels (942 ± 863 pg/mL vs 385 ± 368 pg/mL, p = 0.01) and decreased hemoglobin levels (8.42±1.76 vs 10.0±2.16, *p=*0.02) compared to the normal repeat TRV group. There were no significant differences in the other laboratory values between groups.

Several echocardiographic measures of right ventricular size, function and pressure differed between the groups. Patients in the elevated TRV group had decreased tricuspid annular planar systolic excursion (TAPSE) (1.90 ± 0.52 mm vs 2.23 ± 0.43, p = 0.03), increased right ventricle (RV) size (RV middle diameter: 3.27±0.85 vs 2.72±0.78, *p=*0.04) and decreased velocity time integral stroke volume (61.76±15.10 vs 84.36±27.09, *p=*0.01) compared to the normal TRV group.

### Changes in echocardiographic measures of right ventricular function over time

Among all 51 patients, there were no significant changes from incident to repeat echocardiogram in TRV, TAPSE, or tricuspid annular systolic excursion velocity (RVS’) (Table 4). Similarly, there were no differences between incident and repeat echocardiogram in those variables in the elevated repeat TRV cohort. In the normal repeat TRV cohort, there was a significant difference in TRV from incident to repeat echo, 3.07±0.30 to 2.49±0.20 m/s (*p=*0.01). There was no difference in this cohort in TAPSE or RVS’.

### Predictors of elevated TRV

Univariable logistic regression was performed to identify predictors of elevated TRV (**Table 3**). Among all the variables, significant initial predictors included hemoglobin at hospitalization (p=0.02, 95% CI: 0.46–0.91), TAPSE (p=0.05, 95% CI: 0.05–0.93), RV mid-diameter (p=0.05, 95% CI: 1.04–5.52), left ventricle (LV) stroke volume (p=0.01, 95% CI: 0.91–0.98), LV stroke volume index (SVI) (p =0.02, 95% CI: 0.84–0.98), and heart rate at hospitalization (p=0.02, 95% CI: 1.01–1.10) (Table 2).

**Table 2.**
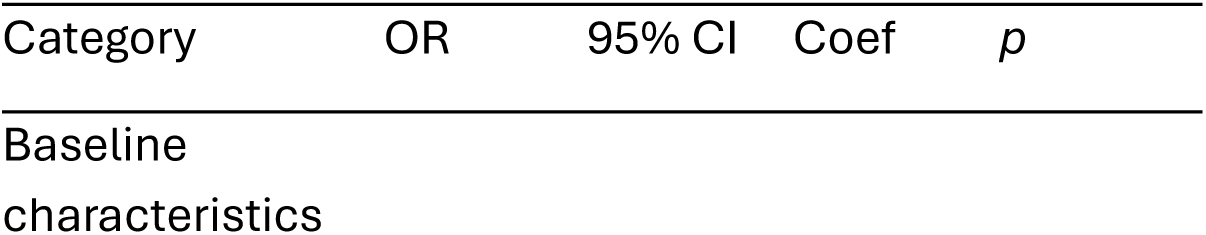
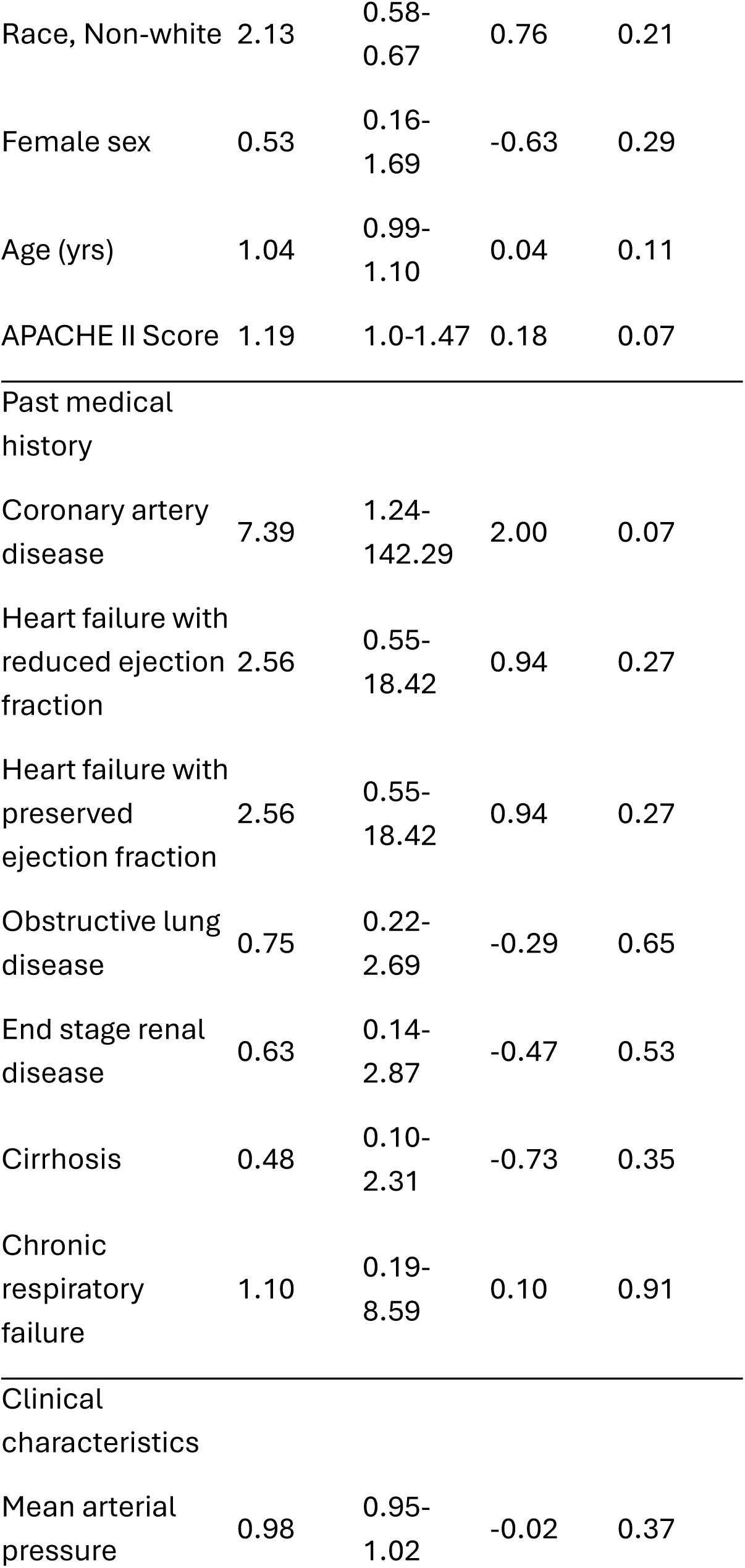
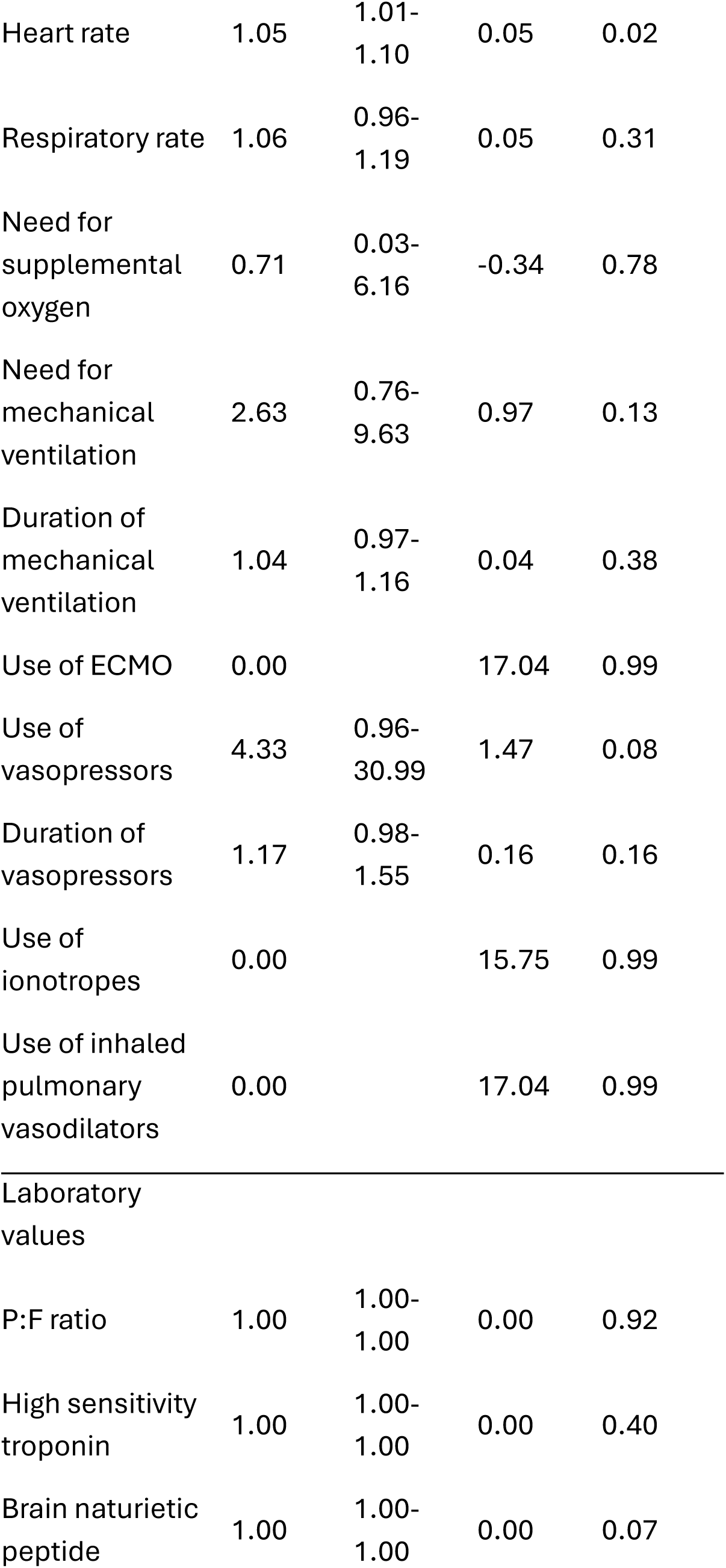
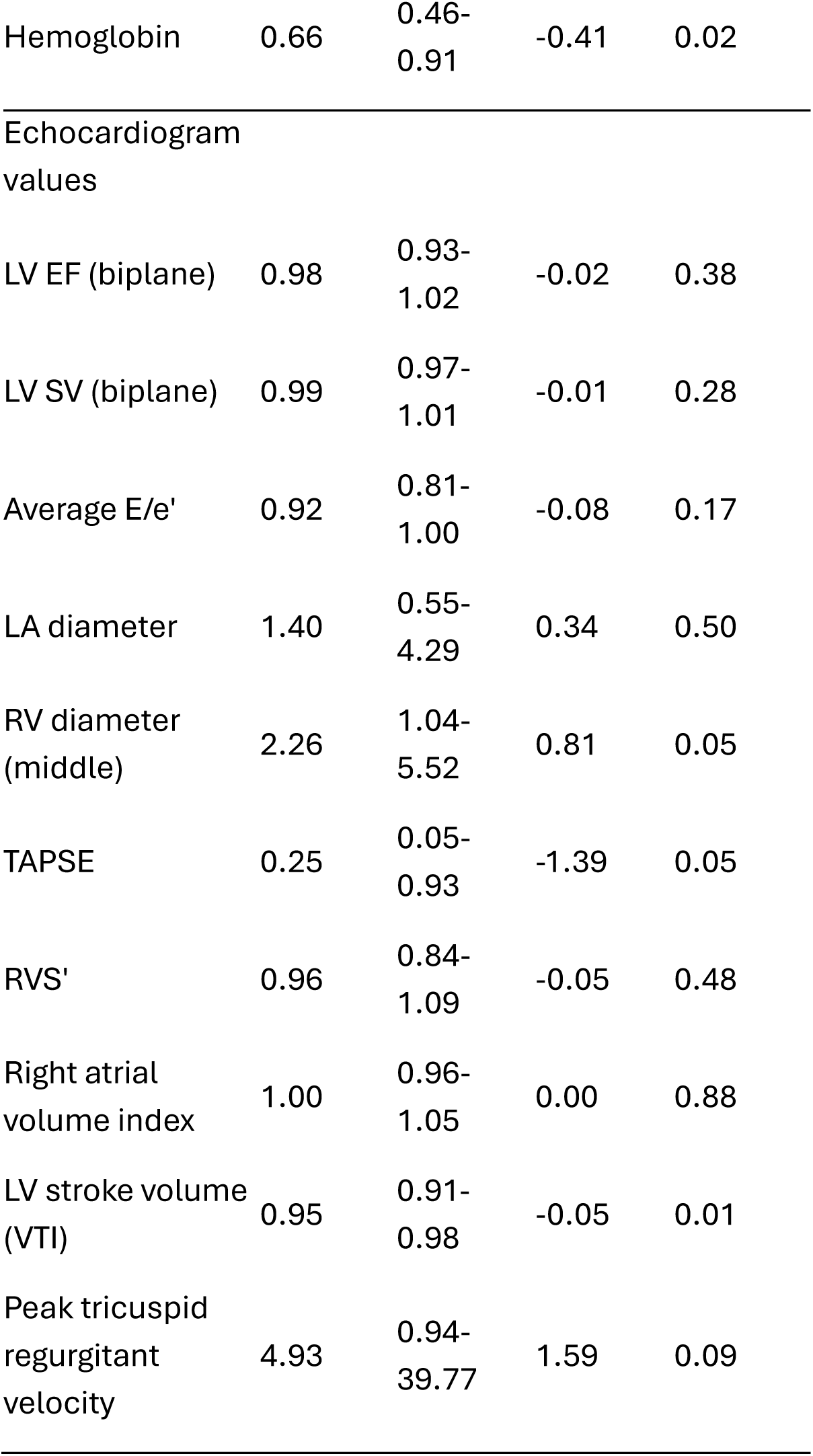
See table 1 legend for expansion of abbreviation. ECMO = extra-corporeal membrane oxygenation; AST = aspartate aminotransferase; ALT = alanine aminotransferase; IVC = inferior vena cava; sPAP = systolic pulmonary arterial pressure; RVOT VTI = right ventricular outflow tract output measured by velocity time integral

| Category | OR | 95% CI | Coef | *p* |
| --- | --- | --- | --- | --- |
| Baseline characteristics |  |  |  |  |
| Race, Non-white | 2.13 | 0.58-0.67 | 0.76 | 0.21 |
| Female sex | 0.53 | 0.16-1.69 | -0.63 | 0.29 |
| Age (yrs) | 1.04 | 0.99-1.10 | 0.04 | 0.11 |
| APACHE II Score | 1.19 | 1.0-1.47 | 0.18 | 0.07 |
| Coronary artery<br>disease | 7.39 | 1.24-142.29 | 2.00 | 0.07 |
| Heart failure with<br>reduced ejection<br>fraction | 2.56 | 0.55-18.42 | 0.94 | 0.27 |
| Heart failure with<br>preserved<br>ejection fraction | 2.56 | 0.55-18.42 | 0.94 | 0.27 |
| Obstructive lung<br>disease | 0.75 | 0.22-2.69 | -0.29 | 0.65 |
| End stage renal<br>disease | 0.63 | 0.14-2.87 | -0.47 | 0.53 |
| Cirrhosis | 0.48 | 0.10-2.31 | -0.73 | 0.35 |
| Chronic<br>respiratory<br>failure | 1.10 | 0.19-8.59 | 0.10 | 0.91 |
| Mean arterial<br>pressure | 0.98 | 0.95-1.02 | -0.02 | 0.37 |
| Heart rate | 1.05 | 1.01-1.10 | 0.05 | 0.02 |
| Respiratory rate | 1.06 | 0.96-1.19 | 0.05 | 0.31 |
| Need for supplemental oxygen | 0.71 | 0.03-6.16 | -0.34 | 0.78 |
| Need for mechanical ventilation | 2.63 | 0.76-9.63 | 0.97 | 0.13 |
| Duration of mechanical ventilation | 1.04 | 0.97-1.16 | 0.04 | 0.38 |
| Use of ECMO | 0.00 |  | 17.04 | 0.99 |
| Use of vasopressors | 4.33 | 0.96-30.99 | 1.47 | 0.08 |
| Duration of vasopressors | 1.17 | 0.98-1.55 | 0.16 | 0.16 |
| Use of inotropes | 0.00 |  | 15.75 | 0.99 |
| Use of inhaled pulmonary vasodilators | 0.00 |  | 17.04 | 0.99 |
| Laboratory values |  |  |  |  |
| P:F ratio | 1.00 | 1.00-1.00 | 0.00 | 0.92 |
| High sensitivity troponin | 1.00 | 1.00-1.00 | 0.00 | 0.40 |
| Brain natriuretic peptide | 1.00 | 1.00-1.00 | 0.00 | 0.07 |
| Hemoglobin | 0.66 | 0.46-0.91 | -0.41 | 0.02 |
| Echocardiogram values |  |  |  |  |
| LV EF (biplane) | 0.98 | 0.93-1.02 | -0.02 | 0.38 |
| LV SV (biplane) | 0.99 | 0.97-1.01 | -0.01 | 0.28 |
| Average E/e' | 0.92 | 0.81-1.00 | -0.08 | 0.17 |
| LA diameter | 1.40 | 0.55-4.29 | 0.34 | 0.50 |
| RV diameter (middle) | 2.26 | 1.04-5.52 | 0.81 | 0.05 |
| TAPSE | 0.25 | 0.05-0.93 | -1.39 | 0.05 |
| RVS' | 0.96 | 0.84-1.09 | -0.05 | 0.48 |
| Right atrial volume index | 1.00 | 0.96-1.05 | 0.00 | 0.88 |
| LV stroke volume (VTI) | 0.95 | 0.91-0.98 | -0.05 | 0.01 |
| Peak tricuspid regurgitant velocity | 4.93 | 0.94-39.77 | 1.59 | 0.09 |

**Table 3.** RV = right ventricle; TAPSE = tricuspid annular plane systolic excursion; SVI (VTI) = stroke volume index by velocity time integral method.

| Category | Coef | OR | 95% CI | P-value |
| --- | --- | --- | --- | --- |
| Intercept | 10.85 | 5.14E+04 | 15.44-3.66E+09 | 0.02 |
| Hemoglobin | -0.57 | 0.64 | 0.30-0.90 | 0.03 |
| RV middle diameter | 1.07 | 2.64 | 0.84-14.65 | 0.13 |
| TAPSE | -1.99 | 0.14 | 0.01-1.08 | 0.09 |
| SVI (VTI) | -0.12 | 0.89 | 0.77-0.98 | 0.03 |

**Table 4.**
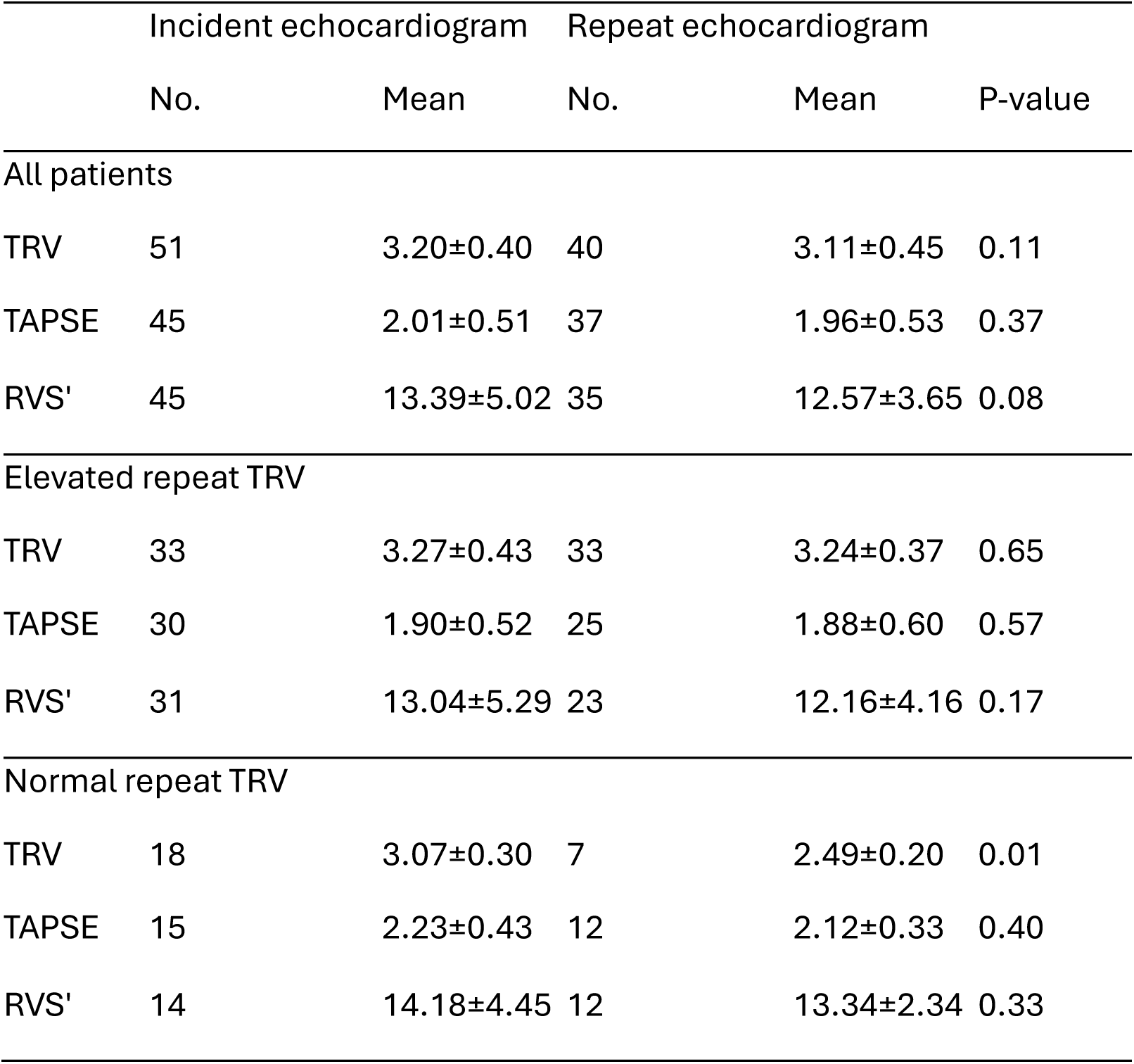
See table 1 legend for expansion of abbreviation.

A final model was created using multivariable logistic regression on the statistically significant univariable predictors. The significant predictors of elevated repeat TRV that remained in the final model included hemoglobin (p=0.03, 95% CI: 0.30–0.90) and SVI (p=0.03, 95% CI: 0.77–0.98). TAPSE and RV middle diameter were not significant in the final model. In addition, elevated SVI (coefficient =0.57) and hemoglobin (coefficient =–0.20) were associated with decreased odds of elevated TRV (**Table 3**).

### Association between TRV cohort and mortality

Kaplan-Meier survival analysis demonstrated a significant association between initial TRV category and mortality. Patients with higher initial TRV had significantly lower survival compared to those with lower TRV values (log-rank test, p=0.046) (**Figure 1**). In contrast, when stratified based on repeat TRV there was no significant difference in survival between patients with repeat TRV> 2.8 and repeat TRV< 2.8 (log-rank test, p=0.33) (**Figure 2**).

**Figure 2.**
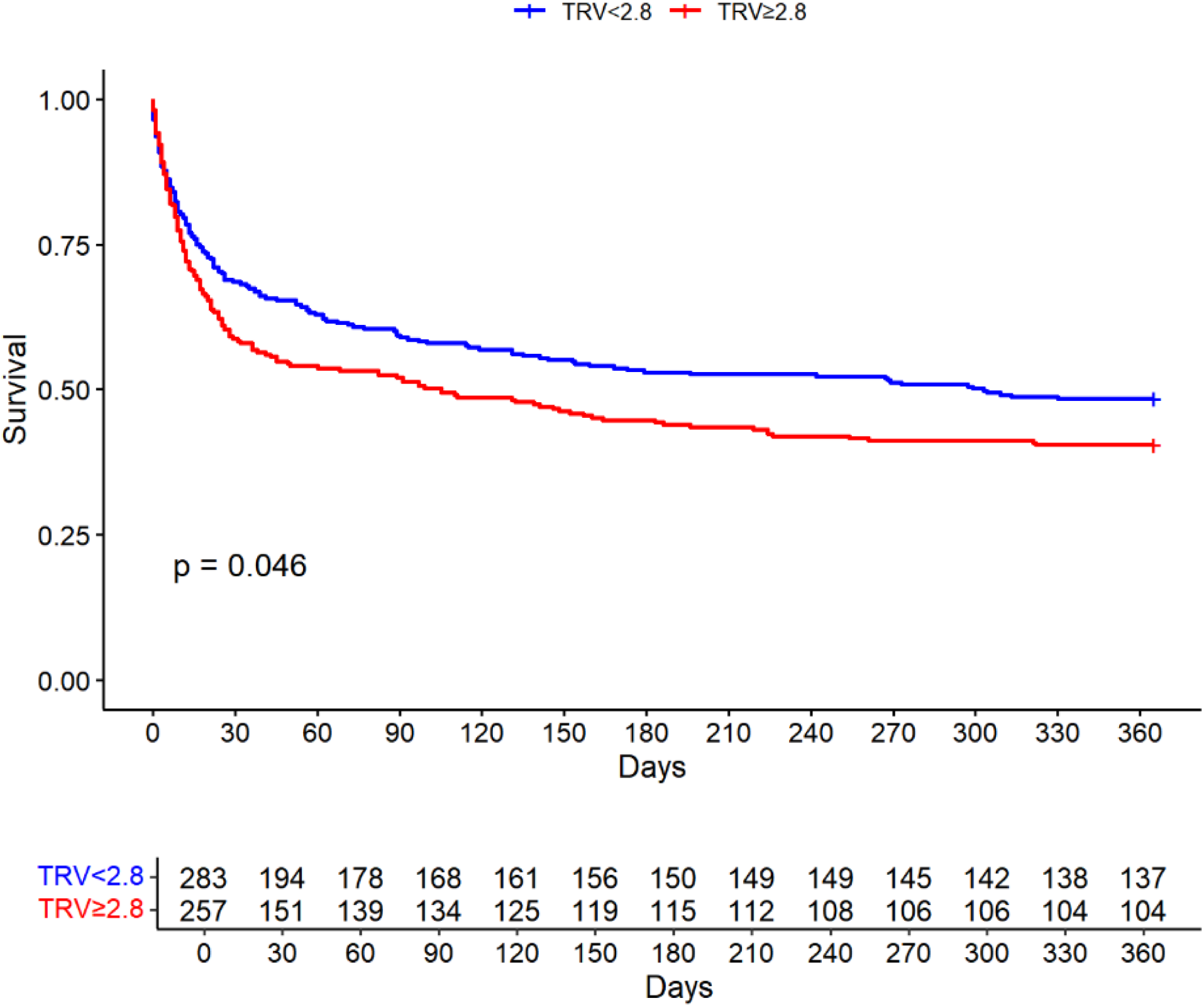
All-cause mortality in patients admitted to the medical intensive care unit at OSUMC in 2021 with a measured TRV. See table 1 legend for expansion of abbreviation.

**Figure 3.**
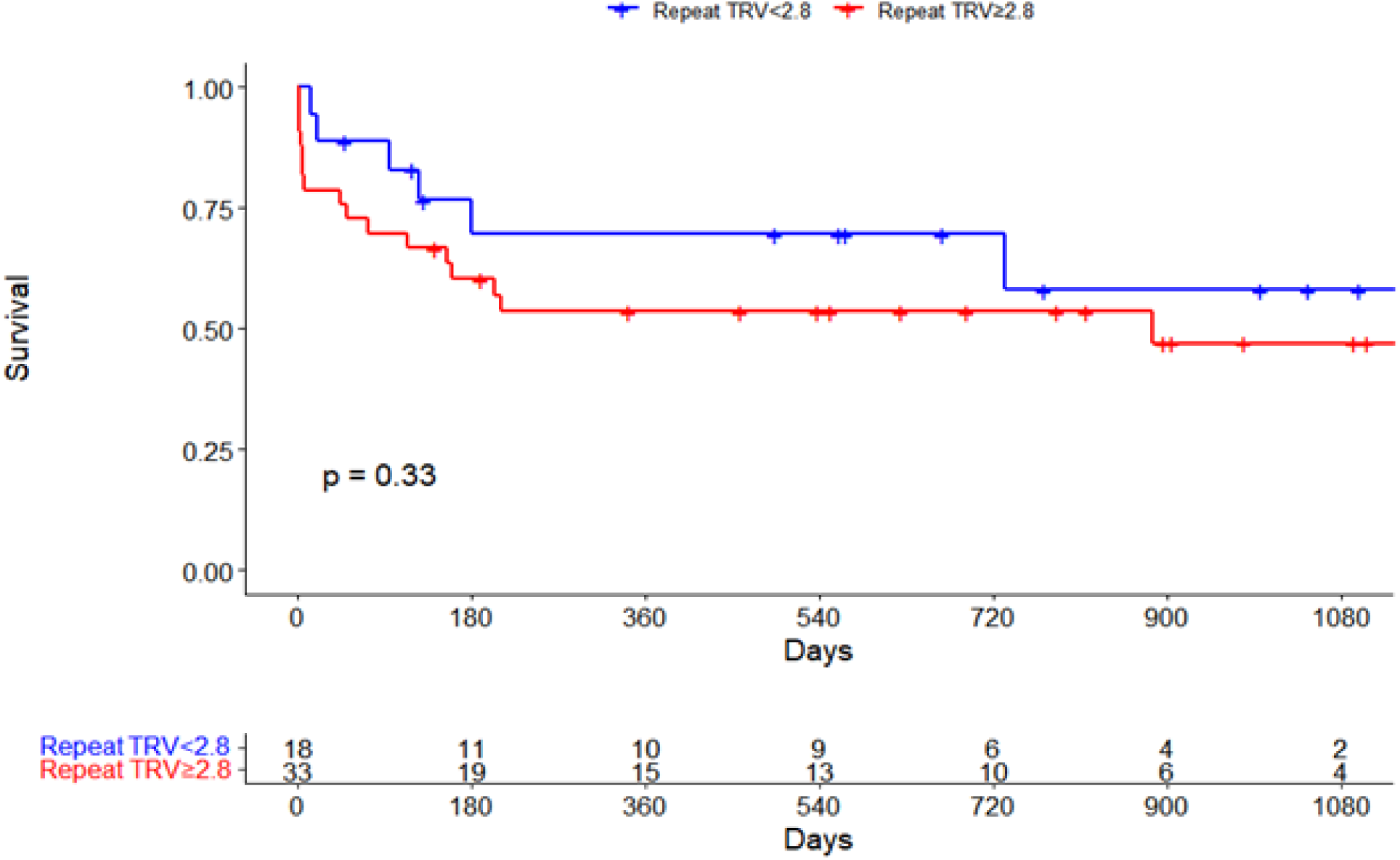
Crude survival after repeat echocardiogram completed after hospital discharge. See table 1 legend for expansion of abbreviations.

**Figure 4.**
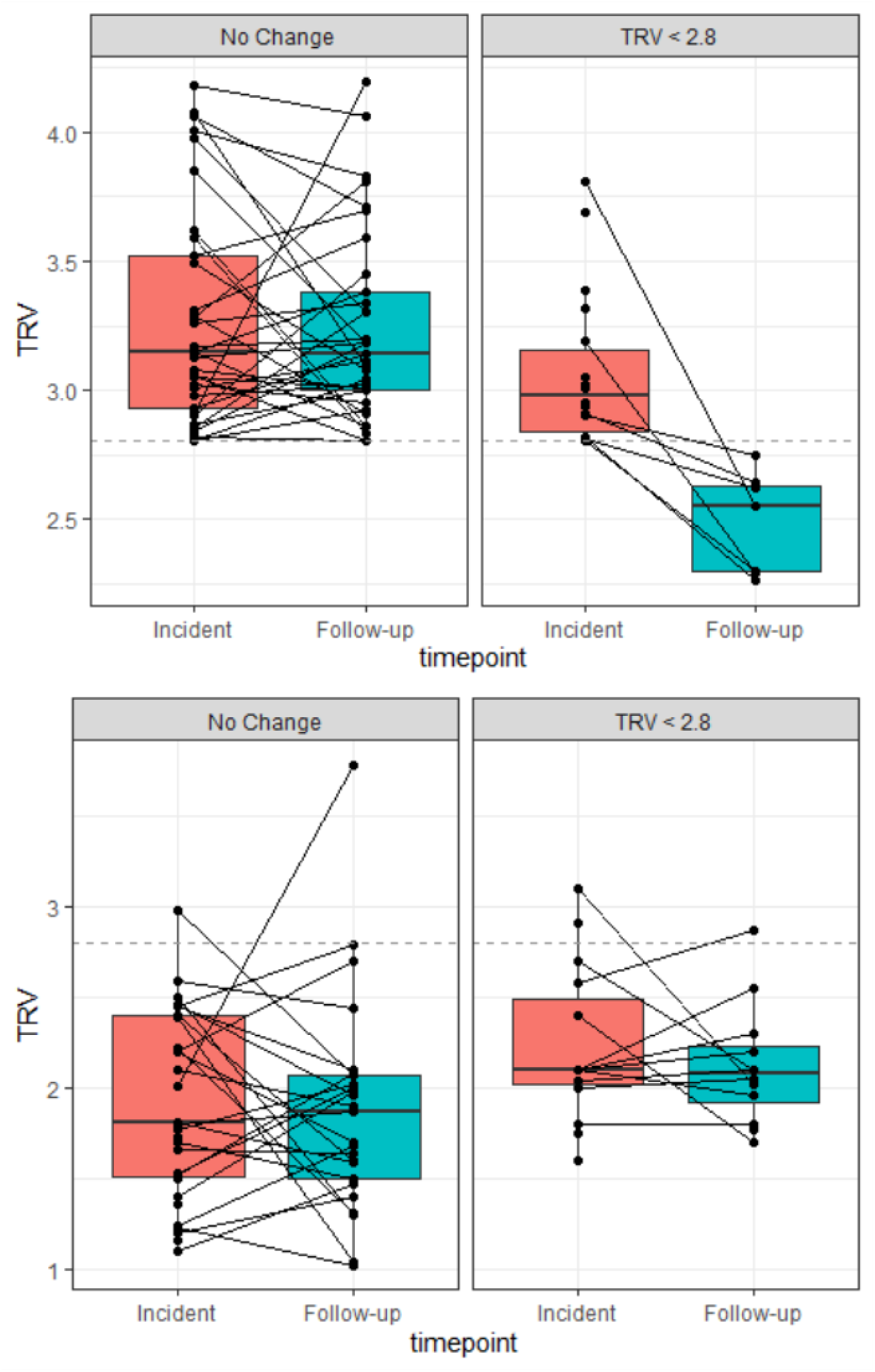
Comparison of right ventricular parameters before and after hospital discharge. A,B,C = all patients with a follow-up echocardiogram. D,E,F = normal repeat TRV cohort. G,H,I = elevated repeat TRV cohort. See table 1 legend for expansion of abbreviations.

### Left ventricular function in critically ill patients

Markers of left ventricular and systolic function on incident echocardiogram were generally similar between the elevated and normal repeat TRV cohorts. Left ventricular ejection fraction was normal in both groups according to the American Society of Echocardiography guidelines, and there was no significant difference between both groups (*p=*0.34) (Table 1)^10^. E/e’ ratio, a marker of left ventricular diastolic function, was elevated in both groups. On average, the E/e’ ratio was higher in the normal repeat TRV cohort (14.41±5.95) than the elevated repeat TRV cohort (12.12±4.89), though this finding did not reach statistical significance (*p=*0.2). Stroke volume by velocity time integral (VTI) was different between groups, with a higher stroke volume in the normal repeat TRV group (84.36±27.09 ml) than the elevated TRV group (61.76±15.10 ml, *p*=0.01).

## DISCUSSION

In this study, we investigated whether elevated pulmonary vascular pressures detected during critical illness persisted after resolution of critical illness. We identified clinical and sonographic variables predictive of persistently elevated pulmonary vascular pressure, and the association of elevated pulmonary vascular pressures with mortality after a follow-up echocardiogram. To our knowledge, this was the first study evaluating echocardiographic markers of right ventricular function over time after critical illness. Inclusion of all ICU patients using real-world data makes our study broadly generalizable, and stimulates further areas of study in subgroups of critically ill patients. Our analysis of many clinical, biochemical, and echocardiographic factors provided a comprehensive assessment of our patients during their critical illness. This study demonstrates that the common clinical scenario regarding often incidentally found elevated pulmonary vascular pressures in critically ill patients warrants further attention after resolution of critical illness and that this area warrants further study,

### Prevalence of elevated pulmonary vascular pressure in critically ill patients

Our study demonstrated a high percentage of critically ill patients (47.6%) have elevated pulmonary vascular pressures, and there is higher mortality in those with elevated pulmonary vascular pressures. This is concordant with previous studies that have primarily been focused on incidence of elevated pulmonary vascular pressures and the impact on mortality during and after the incident hospitalization^1,2,11^. For example, a single center study showed that 42% of all critically ill patients with a recorded TRV had elevated pulmonary vascular pressures as defined by a TRV >3.0 m/s. There was a difference in mortality (37% vs. 25%) between those with and without an elevated TRV. A multivariable logistic regression model identified depressed ejection fraction, low serum bicarbonate, and a diagnosis of pulmonary embolism as predictive variables for elevated TRV^1^. A secondary analysis of the FACTT trial on identified that 92% of intubated patients with acute lung injury had elevated pulmonary vascular pressures as identified by a mean pulmonary arterial pressure on right heart catheterization of >20 mmHg, and that there was an association between both trans-pulmonary gradient and pulmonary vascular resistance index and mortality^2^ A different single center study identified 34.4% of patients with sepsis and septic shock as having doppler defined pulmonary hypertension and that there was a significant difference in one-year mortality between the elevated and normal TRV groups ^11^. These studies demonstrated that elevated pulmonary vascular pressures are prevalent in multiple disease states, and have a significant association with increased mortality. Our findings demonstrate elevated pulmonary vascular pressures during critical illness confirming findings from previous studies and further show elevated pulmonary vascular pressures are likely to persist after resolution of critical illness.

### Study design considerations

We included all patients admitted to the medical intensive care unit at the Ohio State Wexner Medical Center during the year 2021. This approach was chosen as different disease states have been shown to be associated with elevated pulmonary vascular pressures. Additionally, this study is the first of its kind to evaluate persistent elevated pulmonary vascular pressures after resolution of critical illness, and we sought to keep our inclusion criteria as broad as possible. Echocardiogram was chosen as our method of detecting elevated pulmonary vascular pressures as right heart catheterization is rarely done on critically ill patients in our ICU, and echocardiogram is much more likely to have a follow-up study. TRV was our chosen measurement of pulmonary vascular pressure as it is very commonly measured on bedside echocardiogram and is a direct measurement, as opposed to the commonly utilized right ventricular systolic pressure or systolic pulmonary arterial pressure as those require subjective assessment of the inferior vena cava, which is susceptible to transient changes in volume status. The cutoff level of 2.8 m/s was chosen as this was the cutoff velocity for an intermediate probability of pulmonary hypertension by the most recent guidelines published by the European Respiratory Society.^8^

### Markers of left heart dysfunction on persistently elevated TRV

The most common type of PH is World Symposium of PH (WSPH) group 2 or pulmonary hypertension secondary to diseases of the left heart. In our data, the most commonly used echocardiographic measurement of diastolic function, E/e’ ratio, was not different at baseline between the two groups at baseline and were not predictive of elevated repeat TRV on repeat assessment (Table 3). The average E/e’ was, however, elevated (>10) in both groups. While diastolic dysfunction is common and can lead to elevated pulmonary vascular pressures, it does not appear to be more common in patients with persistently elevated pulmonary vascular pressures in this cohort. LV ejection fraction was similar and, on average, normal in both groups and did not predict persistently elevated TRV. Stroke volume, however, was significantly higher in the normal repeat TRV group, and stroke volume index was predictive of persistently elevated TRV. These are seemingly discordant findings regarding measurements of LV systolic function and their impact on pulmonary vascular pressures. One possible interpretation of this is that there is a degree of obstructive shock with in those with elevated pulmonary vascular pressures, and if the increased pulmonary vascular pressure and right ventricular size increases to a degree that impacts the stroke volume, it represents a severity of pulmonary hypertension that is more likely to persist.

### Markers of right heart dysfunction on persistently elevated TRV

Among all measured echocardiographic variables of right heart function, only TRV, TAPSE, and RV middle diameter were different between the two groups, and none were predictive of a persistently elevated TRV in a multivariable logistic regression model. RV middle diameter was predictive in a univariable model, supporting the hypothesis that there is a degree of obstructive shock that indicates the severity of pulmonary hypertension, though based on the multivariable regression model, the RV size is a less impactful measurement than stroke volume index. The difference in TAPSE, a marker of RV systolic function, between the two groups and its usefulness in a univariable regression model, but not the multivariable regression model, additionally supports the hypothesis that right ventricular dysfunction may only impact the persistence of elevated pulmonary vascular pressures if it also impacts left ventricular function. Interestingly, two markers of right ventricular systolic function, TAPSE and RVS’, were normal on incident echocardiogram by the standards of the American Society of Echocardiography despite the elevated pulmonary vascular pressure.

### Biochemical predictors of persistently elevated TRV

The hemoglobin levels differing between the two groups and being predictive of persistently elevated TRV was not anticipated. One hypothesis regarding this difference is that anemia could be driving a high cardiac output state that persists after resolution of critical illness, this hypothesis is further supported by the differences in heart rate on presentation between the two groups. BNP, a hormone secreted by cardiomyocytes in response to ventricular stretch, was elevated in both groups and was higher in the persistently elevated TRV group, but was not predictive of persistently elevated TRV in neither the univariable or multivariable regression models, suggesting that it is not useful as predictor of persistent right ventricular dysfunction, likely due to its rapid degradation and clearance after resolution of ventricular stretch^12^.

### Changes in right ventricular function over time

Among all critically ill patients with a repeat echocardiogram after resolution of critical illness, no markers of right ventricular function including TRV, TAPSE, and RVS’ were different between the incident and repeat echocardiograms. This persists when breaking down the patients into normal and elevated repeat TRV groups, with the exception of the TRV. Right ventricular systolic function appears normal and stable over time despite critical illness and elevated pulmonary vascular pressures. This stability in right ventricular function combined with the stable mortality between the elevated and normal TRV cohorts after repeat assessment is interesting as elevated pulmonary vascular pressures are implicated in the development of right ventricular failure and have been shown to impact mortality in the acute setting^1,2,11,13^. It is unclear why this did not correlate to the post-acute setting in our study and this warrants further investigation.

### Limitations

We acknowledge several limitations to this study. This was a retrospective and single-site study, and requires validation in other institutions. We were unable to control the setting and timing in which repeat echocardiograms were performed, and they may have been performed during a different time of critical illness. Our method of acquiring data required collection of echocardiograms from a large number of patients (n=850) and only a small number (n=51) met our inclusion criteria of having an elevated initial TRV and a repeat assessment after ICU discharge. Potentially reducing the power of this study and increasing the chance of type II error. As such, we are likely underselecting variables that predict persistently elevated pulmonary vascular pressures. Additionally, we may have been unable to detect a difference in mortality between the normal and elevated repeat TRV cohorts. Our sole use of echocardiography to measure pulmonary vascular pressures and precludes us from classifying patients as having pulmonary hypertension, and from classifying patients into WHO groups.

### Conclusions

Persistently elevated pulmonary vascular pressures were common among patients after resolution of critical illness, however persistence of elevated pulmonary vascular pressures did not appear to impact mortality. Severity of elevated pulmonary vascular pressures during critical illness to the degree of impacting stroke volume, and a high output state secondary to anemia are hypothesized to contribute to the persistence of elevated pulmonary vascular pressures. Further investigation is needed to confirm these findings.

## Data Availability

All data produced in the present work are contained in the manuscript

## ABBREVIATION LIST

ARDS: Acute respiratory distress syndrome
BNP: Brain natriuretic peptide
EF: Ejection fraction
E/e’: Peak early mitral inflow velocity / early diastolic mitral annular velocity
HFpEF: Heart failure with preserved ejection fraction
HFrEF: Heart failure with reduced ejection fraction
ICU: Intensive care unit
LA: Left atrium
LV: Left ventricle
PAH: Pulmonary arterial hypertension
RA: Right atrium
RHC: Right heart catheterization
RV: Right ventricle
RV S’: Tricuspid annular systolic excursion velocity
SV: Stroke volume
SVI: Stroke volume index
TAPSE: Tricuspid annular systolic planar excursion
TRV: Tricuspid regurgitant velocity
VTI: Velocity time integral

